# Determinants of SARS-CoV-2 receptor gene expression in upper and lower airways

**DOI:** 10.1101/2020.08.31.20169946

**Authors:** H. Aliee, F. Massip, C. Qi, M. Stella de Biase, J. van Nijnatten, E.T.G. Kersten, N. Z. Kermani, B. Khuder, J. M Vonk, R.C H Vermeulen, U-BIOPRED study group, Cambridge Lung Cancer Early Detection Programme, INER-Ciencias Mexican Lung Program, NHLBI LungMAP Consortium, M. Neighbors, G. W. Tew, M. Grimbaldeston, N. H. T. ten Hacken, S. Hu, Y. Guo, X. Zhang, K. Sun, P.S. Hiemstra, B.A. Ponder, M. J. Mäkelä, K. Malmström, R.C Rintoul, P.A. Reyfman, F.J. Theis, C.A. Brandsma, I. M. Adcock, W. Timens, C.J. Xu, M. van den Berge, R. F. Schwarz, G. H. Koppelman, M.C. Nawijn, A. Faiz

## Abstract

**Background:** The recent outbreak of the severe acute respiratory syndrome coronavirus-2 (SARS-CoV-2), which causes coronavirus disease 2019 (COVID-19), has led to a worldwide pandemic. A subset of COVID-19 patients progresses to severe disease, with high mortality and limited treatment options. Detailed knowledge of the expression regulation of genes required for viral entry into respiratory epithelial cells is urgently needed.

**Methods:** Here we assess the expression patterns of genes required for SARS-CoV-2 entry into cells, and their regulation by genetic, epigenetic and environmental factors, throughout the respiratory tract using samples collected from the upper (nasal) and lower airways (bronchi).

**Findings:** Genes encoding viral receptors and activating protease are increased in the nose compared to the bronchi in matched samples and associated with the proportion of secretory epithelial cells in cellular deconvolution analyses. Current or ex-smoking was found to increase expression of these genes only in lower airways, which was associated with a significant increase in the predicted proportion of goblet cells. Both acute and second hand smoke exposure were found to increase ACE2 expression while inhaled corticosteroids decrease ACE2 expression in the lower airways. A strong association of DNA- methylation with ACE2 and TMPRSS2- mRNA expression was identified.

**Interpretation:** Genes associated with SARS-CoV-2 viral entry into cells are high in upper airways, but strongly increased in lower airways by smoke exposure. In contrast, ICS decreases ACE2 expression, indicating that inhaled corticosteroids are unlikely to increase the risk for more severe COVID-19 disease.

**Funding:** This work was supported by a Seed Network grant from the Chan Zuckerberg Initiative to M.C.N. and by the European Union’s H2020 Research and Innovation Program under grant agreement no. 874656 (discovAIR) to M.C.N. U BIOPRED was supported by an Innovative Medicines Initiative Joint Undertaking (No. 115010), resources from the European Union’s Seventh Framework Programme (FP7/2007-2013) and EFPIA companies’ in kind contribution (www.imi.europa.eu). Longfonds Junior Fellowship. We acknowledge the contribution of the whole U-BIOPRED team as listed in the Appendix S1.’ SDB, FM and RFS would like to thank the Helmholtz Association, Germany, for support.” NIH K08HL146943; Parker B. Francis Fellowship; ATS Foundation/Boehringer Ingelheim Pharmaceuticals Inc. Research Fellowship in IPF. RCR is part funded by Cancer Research UK Cambridge Centre and the Cambridge NIHR Biomedical Research Centre. BAP was funded by programme support from Cancer Research UK. The CRUKPAP Study was supported by the CRUK Cambridge Cancer Centre, by the NIHR Cambridge Biomedical Research Centre and by the Cambridge Bioresource. PIAMA was supported by The Netherlands Organization for Health Research and Development; The Netherlands Organization for Scientific Research; The Netherlands Lung Foundation (with methylation studies supported by AF 4.1.14.001); The Netherlands Ministry of Spatial Planning, Housing, and the Environment; and The Netherlands Ministry of Health, Welfare, and Sport. Dr. Qi is supported by a grant from the China Scholarship Council.

## Introduction

Severe acute respiratory syndrome coronavirus-2 (SARS-CoV-2) is a novel beta-coronavirus that was first detected in an outbreak in Wuhan, China, in late 2019, followed by a rapid spread across the world^1,2^. Infection with SARS-CoV-2 causes coronavirus disease 2019 (COVID-19) which was recognized as a global pandemic by WHO in March 2020. Common symptoms of COVID-19 include fever, cough, fatigue, myalgia and diarrhea, with a subset of patients developing severe disease approximately 1 week after initial symptom onset^3,4^. Severe disease is characterized by dyspnea and hypoxemia progressing to acute respiratory distress syndrome (ARDS) and respiratory failure^5^. Most severe COVID-19 patients are lymphopenic and show thrombosis, in particular in small vessels, while disorders of the central or peripheral nervous system, acute cardiac, kidney and liver injury are seen in a subset of the severe COVID-19 cases^6-8^. Risk factors for developing severe disease are older age, as well as a number of pre-existing conditions such as cardiovascular disease, hypertension, diabetes and obesity^3,8,9^. In addition, chronic lung disease as well as active cigarette smoking has been identified as a potential risk factor for disease progression and adverse outcomes in COVID-19^10^.

The cellular and molecular mechanisms for the increased susceptibility for severe COVID-19 are currently incompletely understood. SARS-CoV-2 has been shown to enter the host cell by binding of the viral spike (S) glycoprotein to the angiotensin converting enzyme-2 (ACE2) on the cell surface^11^, with a slightly higher affinity compared to SARS-CoV^12^. Alternative receptors have also been reported, including CD147(BSG), which has a lower affinity interaction with the S protein^13^, and NPR1^14,15^. Upon S protein binding to the ACE2 receptor, proteolytic cleavage of the S protein by proteases such as TMPRSS2, FURIN and CTSL is required for membrane fusion and cell entry of the virus^11,16^. Expression of these cell entry factors therefore identifies the cells that can be infected with SARS-CoV-2 upon exposure to the virus. Consequently, differences in gene expression of these cell-entry factors as a function of genetics, age, smoking and disease status might contribute to increased susceptibility to viral infection and severe COVID-19^17^. Analysis of gene expression patterns within the Human Cell Atlas consortium^18^ in a range of tissues using single-cell RNA sequencing has identified co-expression of ACE2 and TMPRSS2 in epithelial cells in the conjunctiva of the eye, the upper respiratory tract and the gastro-intestinal tract, with limited expression in the lower respiratory tract and the lung parenchyma^19-21^. Recent analyses within the Human Protein Atlas largely confirmed these RNA data, but also revealed that protein expression of the ACE2 receptor is very low in the upper airways and almost absent from the lower airways and lung parenchyma^22^. These data identify the epithelial cells of the upper airways as the most likely site of entry for SARS-CoV-2 into the respiratory tract, but fail to explain the differences in susceptibility for COVID-19 between different age groups, after cigarette smoke exposure or in patients with pre-existing lung disease. Moreover, the (epi)genetic regulation of the expression of these viral entry genes remains largely uncharacterized. We hypothesize that cigarette smoking, increasing age and pre-existing lung disease all increase the expression of the SARS-CoV-2 cell entry factors, thereby enhancing the susceptibility for COVID-19. To test our hypothesis, we performed a comprehensive analysis gene expression of ACE2, TMPRSS2, CTSL, BSG, NRP1 and FURIN in upper and lower airway samples as a function of age, smoking history and disease, as well as the association with SNPs and CpG methylation.

## Materials and Methods

Complete methods, including description of study populations, sample management as well as generation of genomic data are described in the online supplement.

### Gene expression analysis

Differential gene expression analysis was conducted across seven different cohort studies (Table 1) using the R packages EdgeR and Limma for RNA-Seq and microarray studies, respectively. Extensive details about each study can be seen in the online Supplement. All analyses were corrected for age, sex and smoking status. Paired analyses were conducted using limma blocking or EdgeR GLM models correcting for patient ID in microarray and RNA-Seq datasets, respectively. All analyses were conducted as candidate gene functions focusing on *ACE2, TMPRSS2, BSG FURIN, NRP1* and *CTSL*.

**Table 1.** Clinical characteristics of datasets.

| Study | NORM |  | CRUKPAP |  | UBIOPRED |  | PIAMA | Finnish | TIP | OLIVA | GLUCOLD | Mexico City | INDURAIN |
| --- | --- | --- | --- | --- | --- | --- | --- | --- | --- | --- | --- | --- | --- |
| Samples type | nasal brush | bronchial brush | nasal scoop | bronchial brush | nasal brush | bronchial biopsy | nasal brush | bronchial biopsy | bronchial brush | nasal brush | bronchial biopsy | nasal curettage | bronchial biopsy |
| n | 92 | 92 | 405 | 228 | 89 | 108 | 325 | 22 | 79 | 57 | 56 | 79 | 184 |
| age mean (SD) | 36.4(16.6) | 39.3(17.2) | 65.3 (10.7) | 67.4 (9.9) | 43.0(14.9) | 45.1(14.2) | 16.4 (0.2) | 1.1(0.6) | 41.1(18.5) | 45.7(11.9) | 60.6(7.7) | 57.2 (18.5) | 44.1(14.9) |
| sex male n(%) | 49(53.2) | 52(56.5) | 249 (61.5) | 155 (68.0) | 46(51.7) | 54(50) | 141 (43.4) | 20(90.9) | 61(77.2) | 27(47.4) | 50(89.3) | 21 (26.6) | 95(51.6) |
| smoking status<br>current smoking<br>n(%) | 48(52.1) | 45(48.9) | 122 (30.1) | 77 (33.8) | 68(76.4) | 82(75.9) | 23 (7.1) | 0(0) | 77(100) | 27(47.4) | 38(67.9) | 26(33.0) | 59(32.1) |
| Platform | Microarray,<br>RNA Seq | Microarray | RNA Seq | RNA Seq | Microarray | Microarray | RNA Seq | RNA Seq | Microarray | RNA Seq | RNA Seq | RNA Seq | RNA Seq |

### Expression Quantitative Trait Loci (eQTLs)

We assessed the association between gene expression of *ACE2, TMPRSS2, BSG FURIN, NRP1* and *CTSL* and SNPs located within 1MB from the gene body, using the r package matrixeQTL, correcting for age, sex and smoking status (and batch effect in the CRUKPAP study). Meta-analysis of cis-eQTL analysis was performed for Nasal brush: CRUKPAP (n=339), NORM (n=92) and PIAMA (n=303) and Bronchial brush CRUKPAP (n=215) and NORM/TIP (n=150).

### Expression Quantitative Trait Methylation (eQTMs)

We performed cis-eQTM analysis, with a window of +/-1 MB from the gene body. The methylation M value (log2 ratio of methylated versus unmethylated probe intensities) was regressed on gene expression and adjusted for age, sex, and smoking status using the r package matrixeQTL, correcting for age, sex and smoking status. EQTM analysis was conducted on two tissue types; Nasal;- OLIVIA (n=50) and PIAMA (n=245); Bronchial biopsy; INDURAIN (n=144) and GLUCOLD (n=55).

### Meta-analysis

The weighted Z-score method was used to meta-analyze the results of association analysis and eQTL analysis from all cohorts, considering that different cohorts may use different platforms in measuring gene expression data.

### Cellular deconvolution using single-cell RNA sequencing (scRNA-Seq) signatures

scRNA-Seq signatures from the nose and bronchus were utilized from our previously-published data to estimate differences in cell-type composition, using mRNA expression levels^23^. Cellular deconvolution is explained in detail in the supplementary materials.

## Results

### SARS-CoV-2 cell entry genes are increased in expression in the nose compared to the bronchus

We first investigated the expression of *ACE2, TMPRSS2, BSG, FURIN, NRP1* and *CTSL* in matched nasal and bronchial brushings in healthy individuals (NORM, n=77) and a second cohort of patients with suspected cancer (CRUKPAP, n=162). Expression of all six genes was higher in nasal compared to bronchial brushings in both independent datasets (Table 2 and Figure 1A-D). We next investigated the influence of smoking status, age, BMI and sex on COVID related gene expression. A meta-analysis of the two datasets shows that smoking significantly upregulates *ACE2, TMPRSS2, FURIN*, and *BSG* in bronchial but not in nasal brushes (Figure 1E&F). In contrast, smoking was associated with lower expression of *CTSL* in both nasal and bronchial brushings (Table 3). Sex had a small, but significant effect on *BSG* expression only in nasal brushes, while no effects were seen for age (Table S1&2). We next investigated the effect of smoking cessation on *ACE2* expression. We observed significantly decreased bronchial *ACE2* expression as soon as after 1 month of smoking cessation, within expression levels decreasing to the level of never smokers after 12 months (Figure 1G).

**Table 2.**
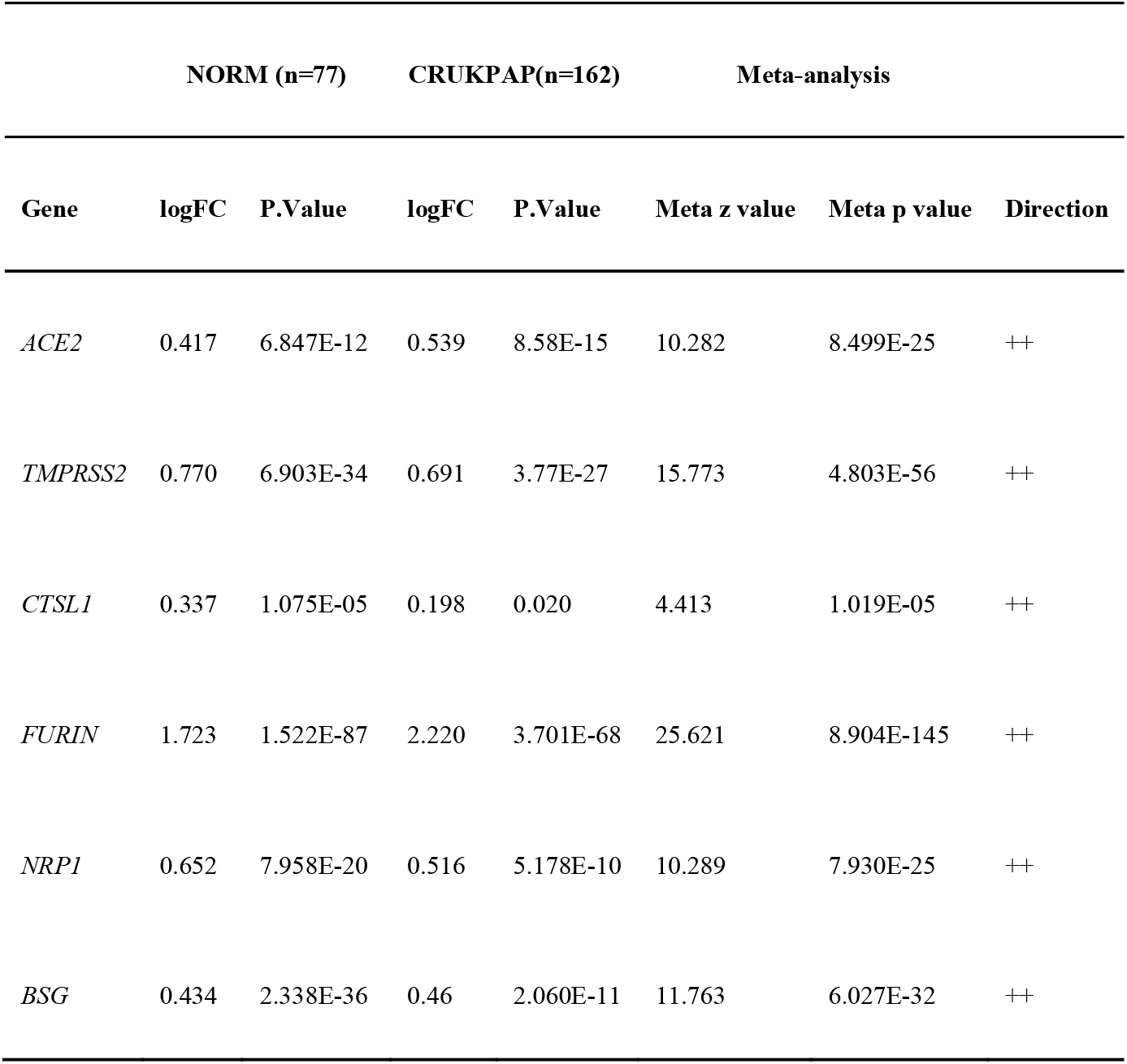
Comparison of nasal and bronchial brushes for COVID-19 related genes in match samples.

**Figure 1.**
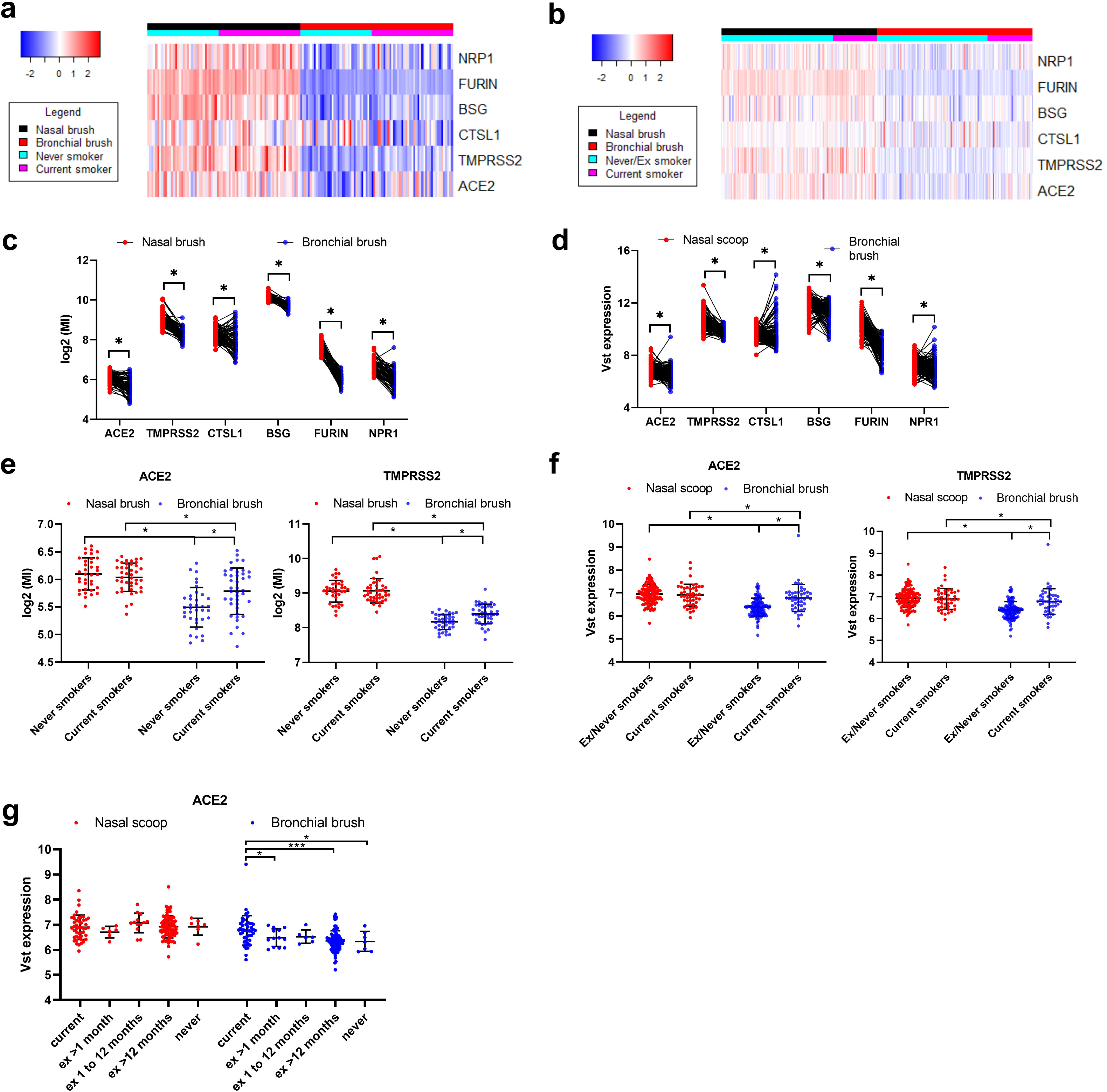
Expression of COVID related genes in nasal and bronchial brushes and relationship with goblets cells. Heatmaps and plots of COVID related genes in matched nasal and bronchial brushes A&C) NORM (n=77) and B&D) CRUKPAP (n=162). Plots comparing ACE2 and TMPRSS2 expression in current and ex/never smokers in nasal and bronchial brushes, E) NORM and F) CRUKPAP. Plots comparing ACE2 expression in current and ex/never smokers separated based on duration of smoke cessation in nasal and bronchial brushes, G) CRUKPAP. *=pvalue<0.05, ***=pvalue<0.001 Abbreviations: MI= microarray intensity

We next performed cellular deconvolution to assess whether cellular composition of nasal and bronchial samples was associated with expression of some of these genes. To this end, scRNA-Seq data obtained from bronchial (n=9) and nasal brush (n=1) were used. The UMAP plot of the data shows that goblet and secretory cells as well as ciliated(nasal) and ciliated(bronchial) cells are well separated. Therefore, we proposed to use location-specific annotations (uppers versus lower airways) for feature selection to capture differentiating genes between these cell types. After feature selection, ciliated(nasal) and ciliated(bronchial) were highly similar, so we combined their expression profiles for deconvolution (Figure 2A&B). Cell-type specific analysis revealed that *ACE2* and *TMPRSS2* were mainly found in nasal goblet and ciliated cells with lower expression in the same cell types in the bronchus ^20^, whereas *BSG* was ubiquitously expressed and *NPR1* was mainly expressed in basal epithelial cells (Figure 2C-H). Cellular devolution of ‘bulk’ RNA-seq data from matched nasal and bronchial brushes samples resulted in similar results, with *ACE2, TMPRSS2, BSG, FURIN, NRP1* and *CTSL* expression strongly correlating with predicted goblet cell numbers (Figure 2I&J). *ACE2* specifically was found to be highly correlated with goblet cell proportions indicating possible selective expression in goblet cells (Figure 2F&G), this result is supported by previously published sc-seq and scATAC-seq data from the human cell atlas^24^. As the goblet cell fraction was found to be higher in the nose than bronchus in this and other studies^25^. Finally, we compared the cellular deconvolution between current smokers and never/ex smokers in matched nasal and bronchial samples. Here we showed a significant increase in secretory cells (nasal + bronchial secretory cell signature combined) in bronchial but not nasal samples of current smokers compared to ex/never smokers in one of the two studies, (Figure 2H&I). The lack of a difference in the bronchial samples of the NORM study is likely due to the difference in performing cellular deconvolution with microarray data, which is evident when we focus only on the bronchial secretary cell signature which is significant in both datasets (Figure S1). These findings indicate that the bronchial specific increase of *ACE2* in response to smoke exposure is likely due to an increase in the proportion of secretory cells in the bronchus, which was not observed in nasal samples.

**Figure 2.**
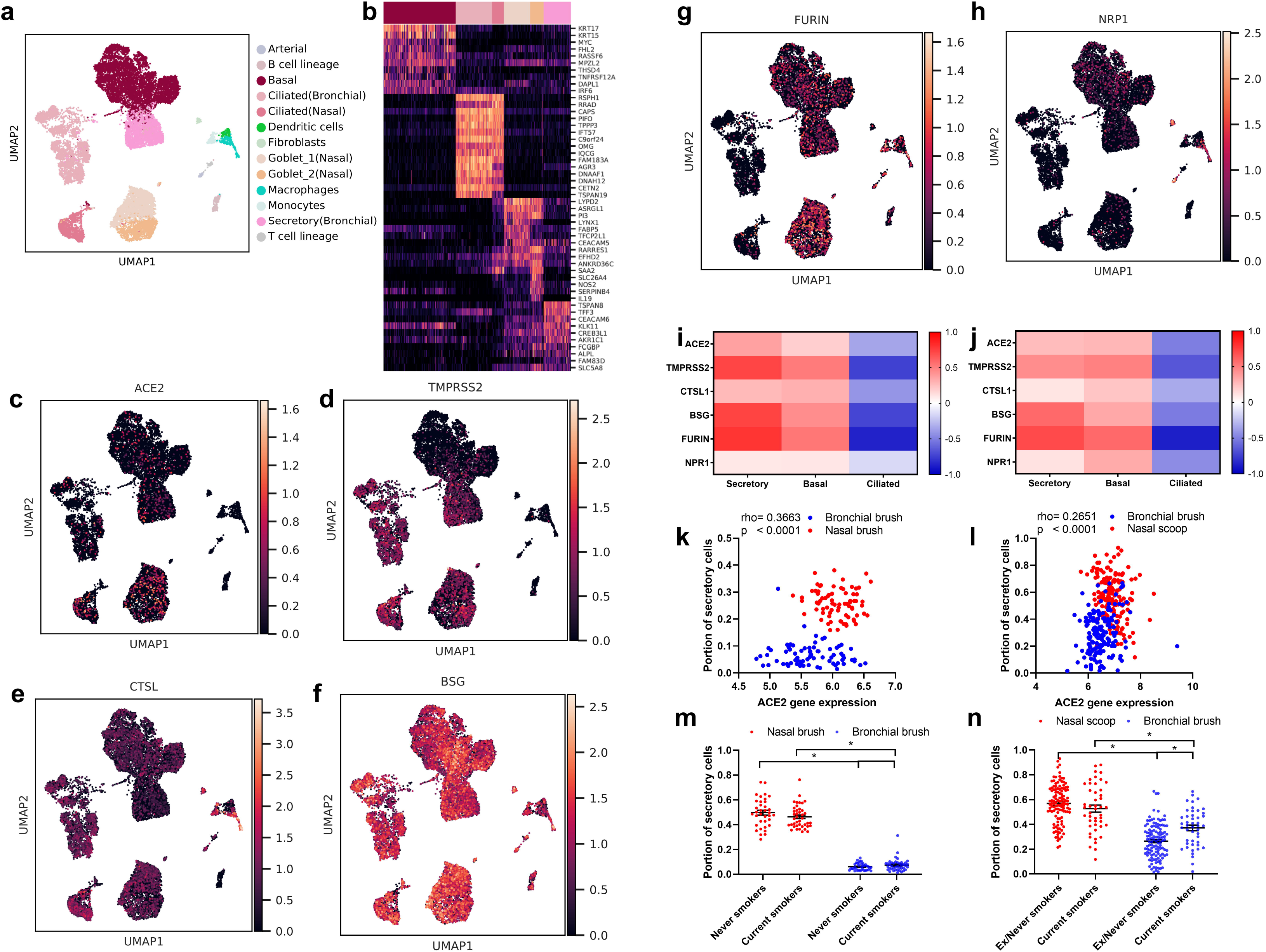
Celular deconvolution of nasal and bronchial samples. a) UMAP of merged bronchial biopsy and nasal brush single cell datasets. b) Heatmap of selected genes associated with each epithelial cell type. UMAP for c) ACE2, d) TMPRSS2, e) CTSL, f) BSG, g) FURIN and h) NRP1. Correlation heatmap of cellular deconvolution cell proportions compared to COVID related genes, i) NORM and j) CRUKPAP. Association of cellular deconvolution of Goblet cells with ACE2 expression, k) NORM and l) CRUKPAP. Goblet/secretory cell fraction separated based on tissue type and smoking status, m) NORM and n) CRUKPAP. Statistics for deconvolution results were conducted using Mann-Whitney test. *=p<0.05

### Relationship of nasal and bronchial genes associated with SARS-CoV-2 entry with smoking, sex, age and BMI

We next expanded our analyses to include larger non-matched cohorts of airway samples. As the nose is thought to be the primary site of COVID infection, we first investigated the relationship between nasal expression of SARS-CoV-2 cell entry genes and factors that have been epidemiologically linked to COVID severity (smoking, sex, age and BMI). We ran our analysis on five populations with RNA-Seq data: adult, NORM/OLIVA (n=76), CRUKPAP dataset (n=405), UBIOPRED (n=89), and INER-Ciencias Mexican Lung Program (INCI) (n=79); children, PIAMA (n=291); a meta-analysis was then run on the five studies. Through meta-analysis *TMPRSS2* expression was found to be significantly increased by current smoking in two of the five cohorts but this finding was inconsistent across all five cohorts, while *CTSL* expression was decreased in all cohorts. In all studies, no association was found with *ACE2* expression in all variables tested (smoking, sex, age and BMI)(Table 4). Overall little association was found for the expression of COVID related genes with epidemiologically linked factors in the nose.

**Table 3.**
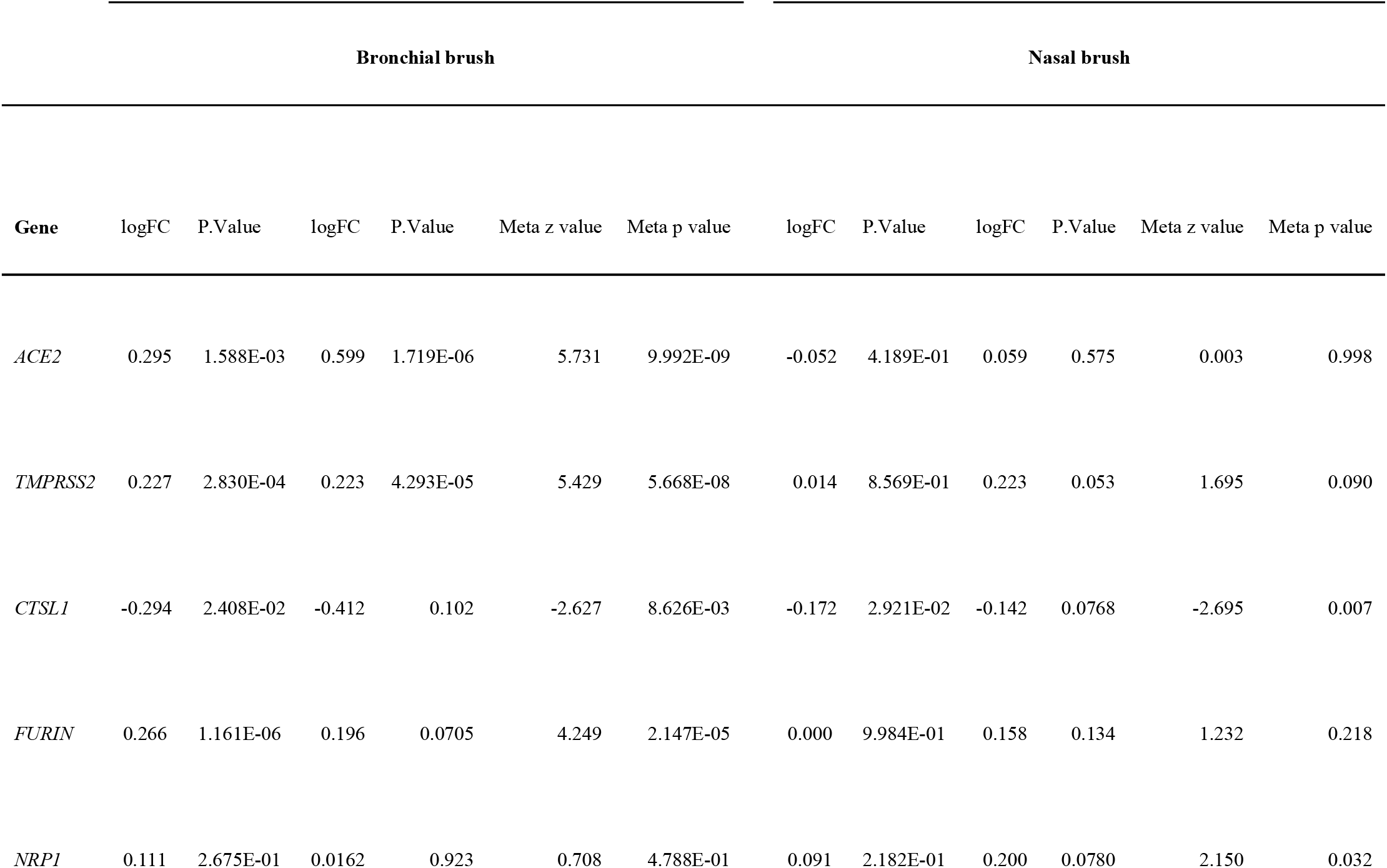
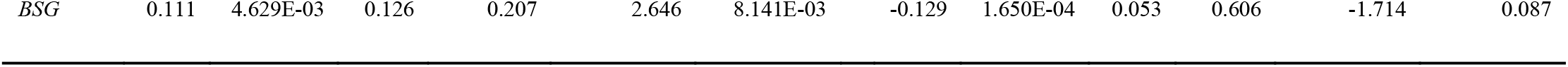
Influence of current smoking on COVID-19 related genes in matched nasal and bronchial brushes.

We next repeated this analysis in samples obtained from bronchial brushes and bronchial (forceps) biopsies. We ran our analysis on five adult populations: bronchial biopsies; INDURAIN (n=184), UBIOPRED (n=108) and GLUCOLD (n=56) and bronchial brushes CRUKPAP dataset (n=228) and NORM/TIP (n=167); a meta-analysis was then run on the bushes and biopsies separately (Table 5). Analysis of these samples shows a clear upregulation of ACE2, TMPRSS2, FURIN, and BSG in current versus never/ex-smokers in both bronchial biopsies and bronchial brushes (Figure 3A-F). While CTSL expression was found to be lower in bronchial biopsies but unchanged in bronchial brushes. Cellular deconvolution of these samples shows an association of these genes with secretory cells across two of four cohorts (Figure 3G).

**Table 4.**
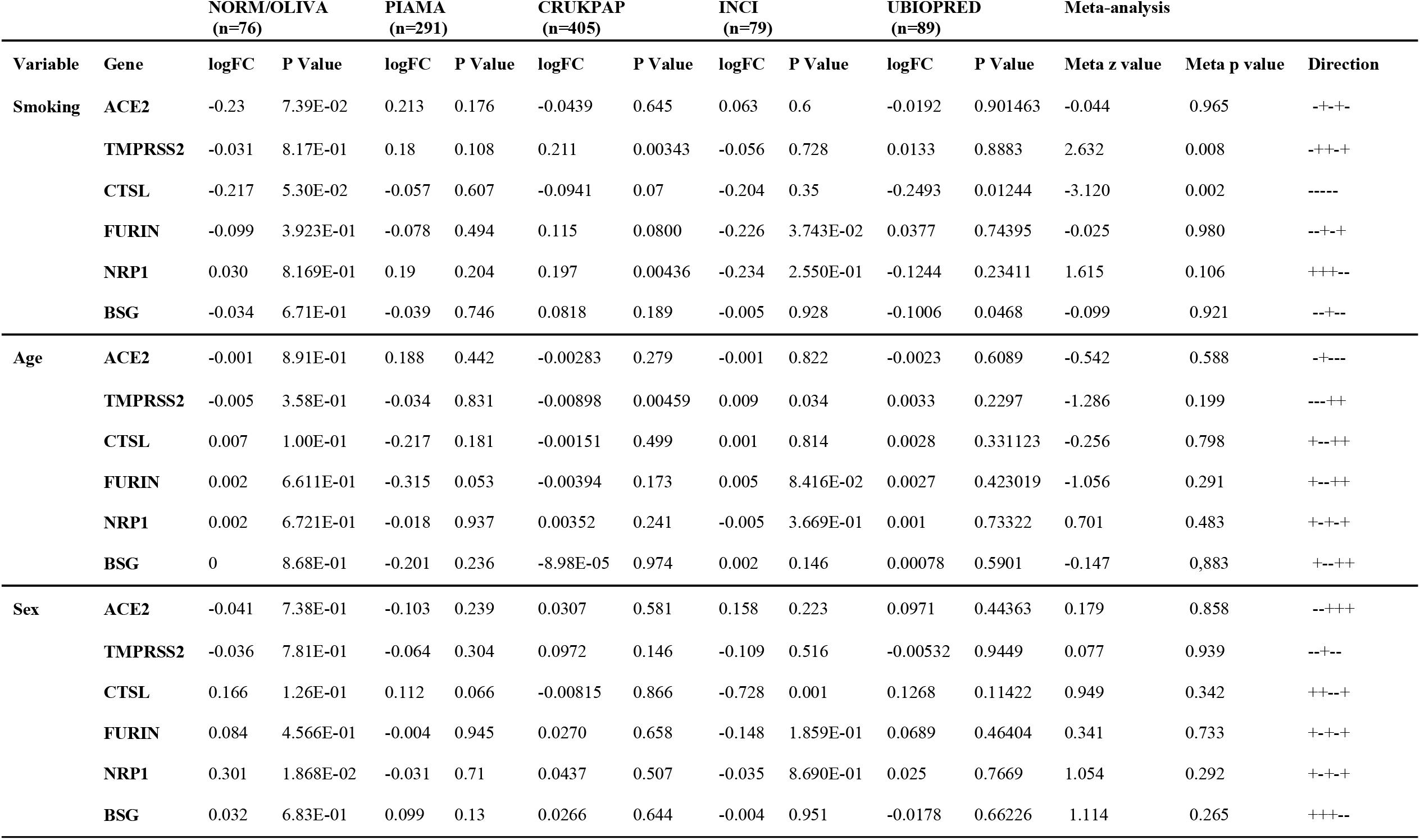
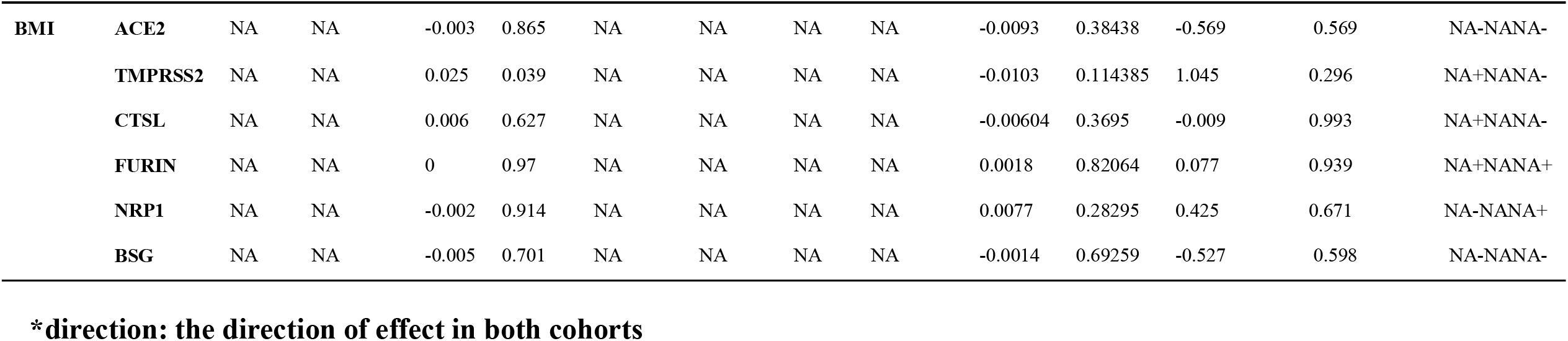
Influence of patient characteristics on COVID related genes measured in nasal brushes.

**Table 5.**
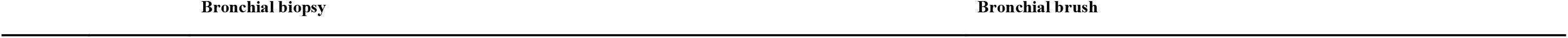
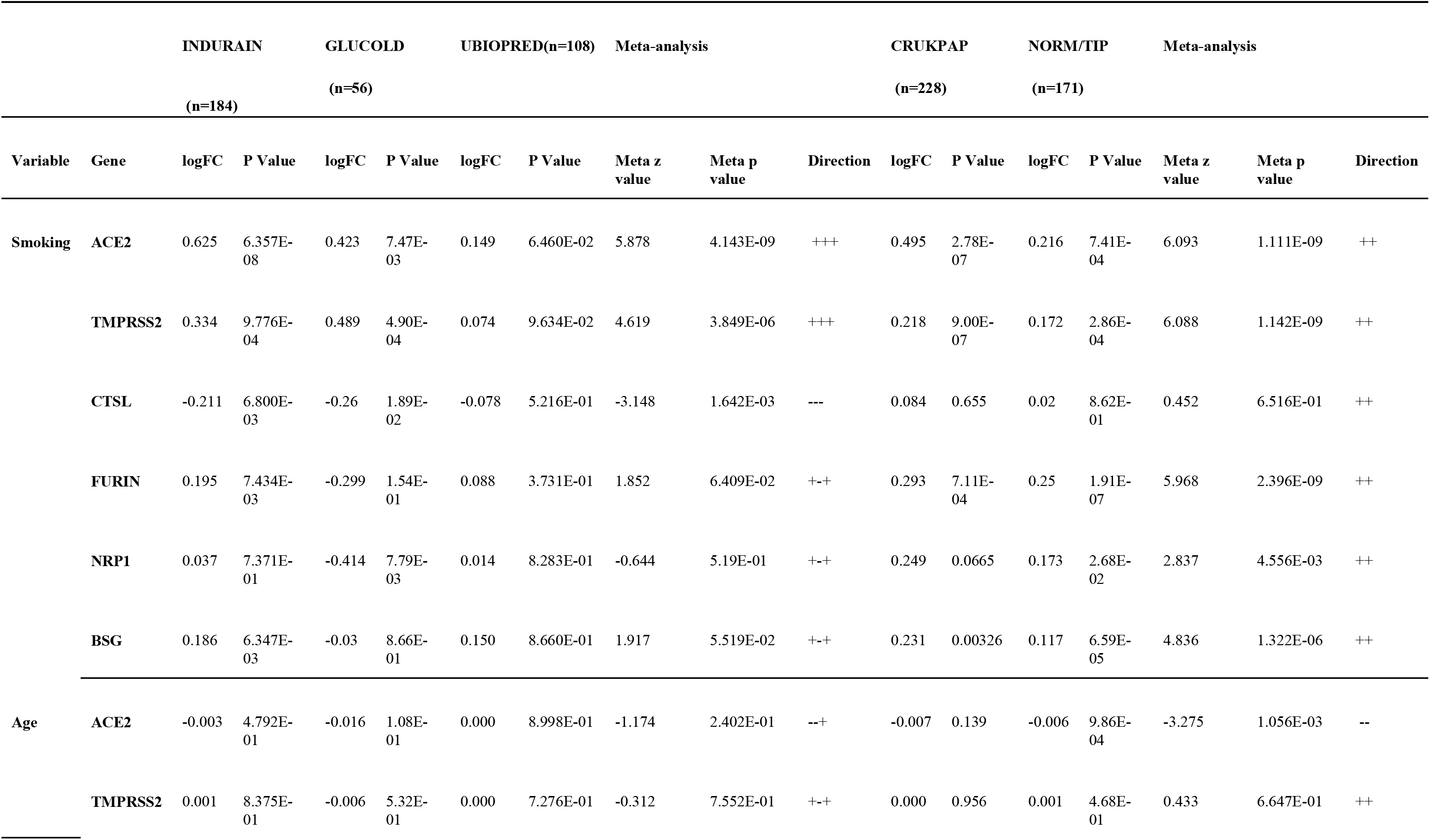
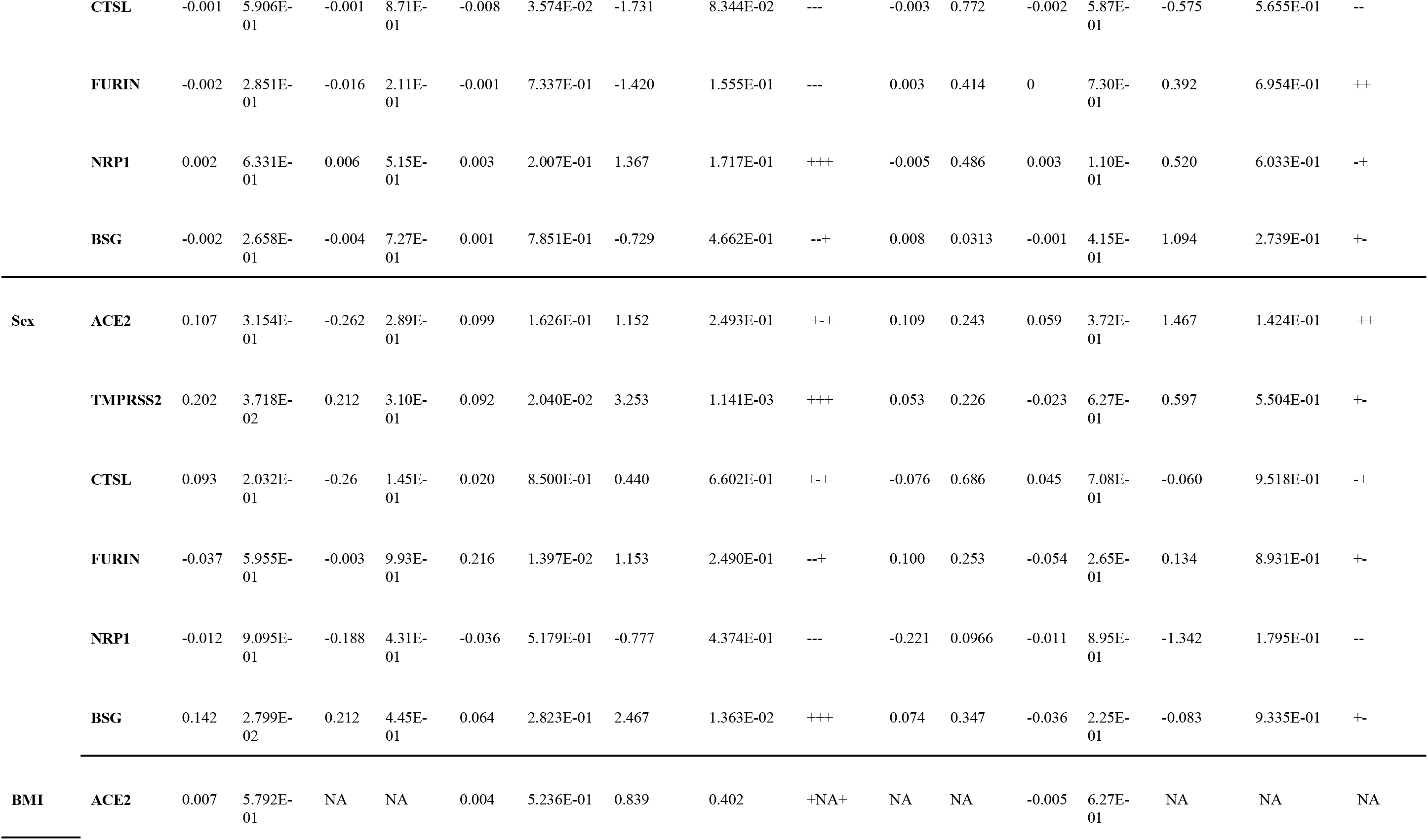
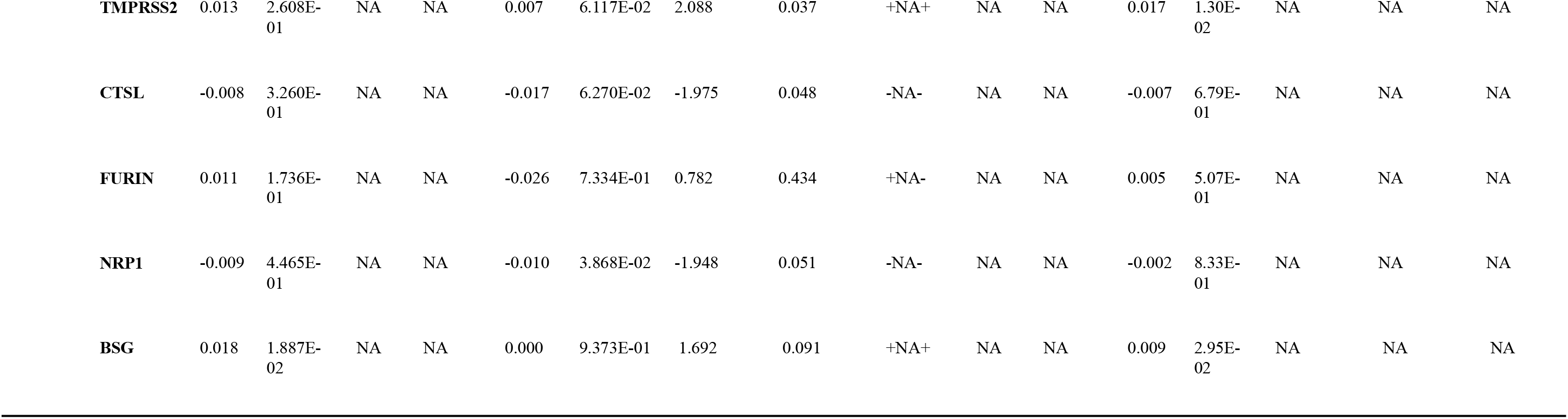
Influence of patient characteristics on COVID related genes measured in bronchial samples.

**Figure 3.**
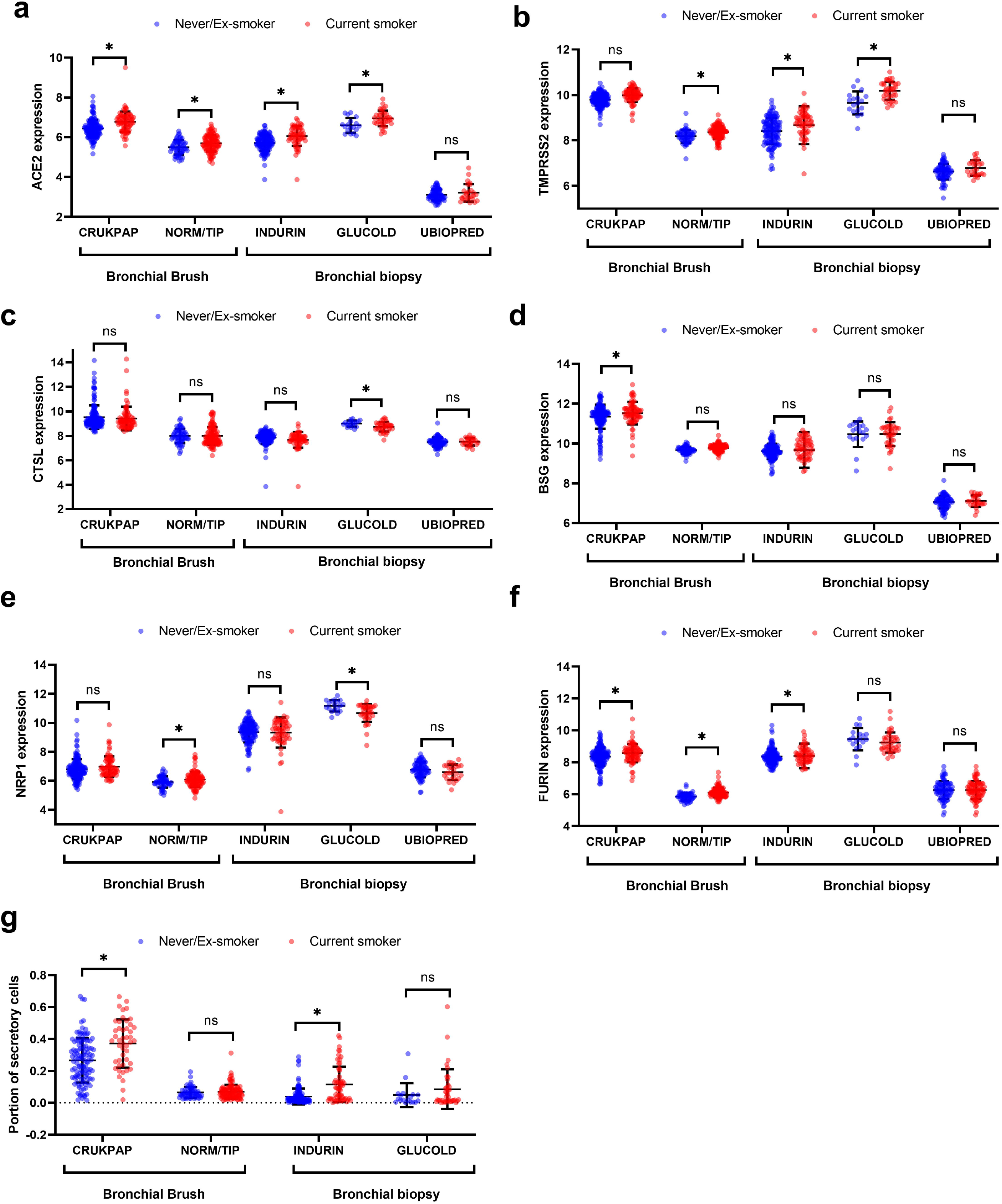
Meta-analysis of COVID related genes in nasal and bronchial brushes. Expression of a) ACE2, b) TMPRSS2, c) CTSL, d) BSG, e) NRP1, f) FURIN in bronchial biopsies; INDURAIN (n=207), UBIOPRED (n=108) and GLUCOLD (n=56) and bronchial brushes CRUKPAP dataset (n=228) and NORM/TIP (n=167), separated based on smoking status. g) cellular deconvolution of bronchial brushing and biopsies separated based on smoking status. Cellular deconvolution was statisitcs was done using a unpaired t-test. *=p<0.05

### Acute smoke exposure and second hand smoke in children is associated with ACE2 expression

Although chronic exposure to smoke increases genes associated with SARS-CoV-2 entry, we next investigated whether acute smoke exposure (24 hours) is sufficient to have a similar effect. Here we investigated “social smokers” who were asked to abstain from smoking for 2 days, followed by either smoking or not smoking of three cigarettes over 1 hour. A bronchial brush was then taken 24 hours later. We saw a significant increase (p=1.029E-02) in *ACE2* expression following acute smoke exposure and a trend towards an increase was found for the alternative receptor *NRP1* (p=6.592E-02) (Figure 4A, Table S3), while no change was seen in the other four genes. These results provide evidence that acute smoking can increase *ACE2* expression without altering structural cellular composition which is unlikely to change over a 24 hours period. We also investigated the effects of second-hand smoke in infants of parents who smoke. In infants (aged 13.5 ± 6.9 months) exposed to second-hand smoke (n=9) compared to controls (n=13), we observed a significantly higher expression of *ACE2* and *FURIN*, while no significant difference in the expression of *CTSL, TMPRSS2, NRP1* and *BSG* (Figure 4B, Table 6). These results indicate that second-hand smoke may be sufficient to increase gene expression in children, however due to the low number of samples in the current analysis these results need to be viewed with caution.

**Table 6.** Influence of second hand smoke on expression of COVID related genes in bronchial biopsies.

| Gene | logFC | PValue |
| --- | --- | --- |
| ACE2 | 0.876 | 4.448E-02 |
| TMPRSS2 | 0.252 | 2.277E-01 |
| CTSL | 0.554 | 6.861E-02 |
| BSG | 0.284 | 1.229E-01 |
| FURIN | 0.616 | 9.946E-04 |
| NRP1 | 0.344 | 2.372E-01 |

**Figure 4.**
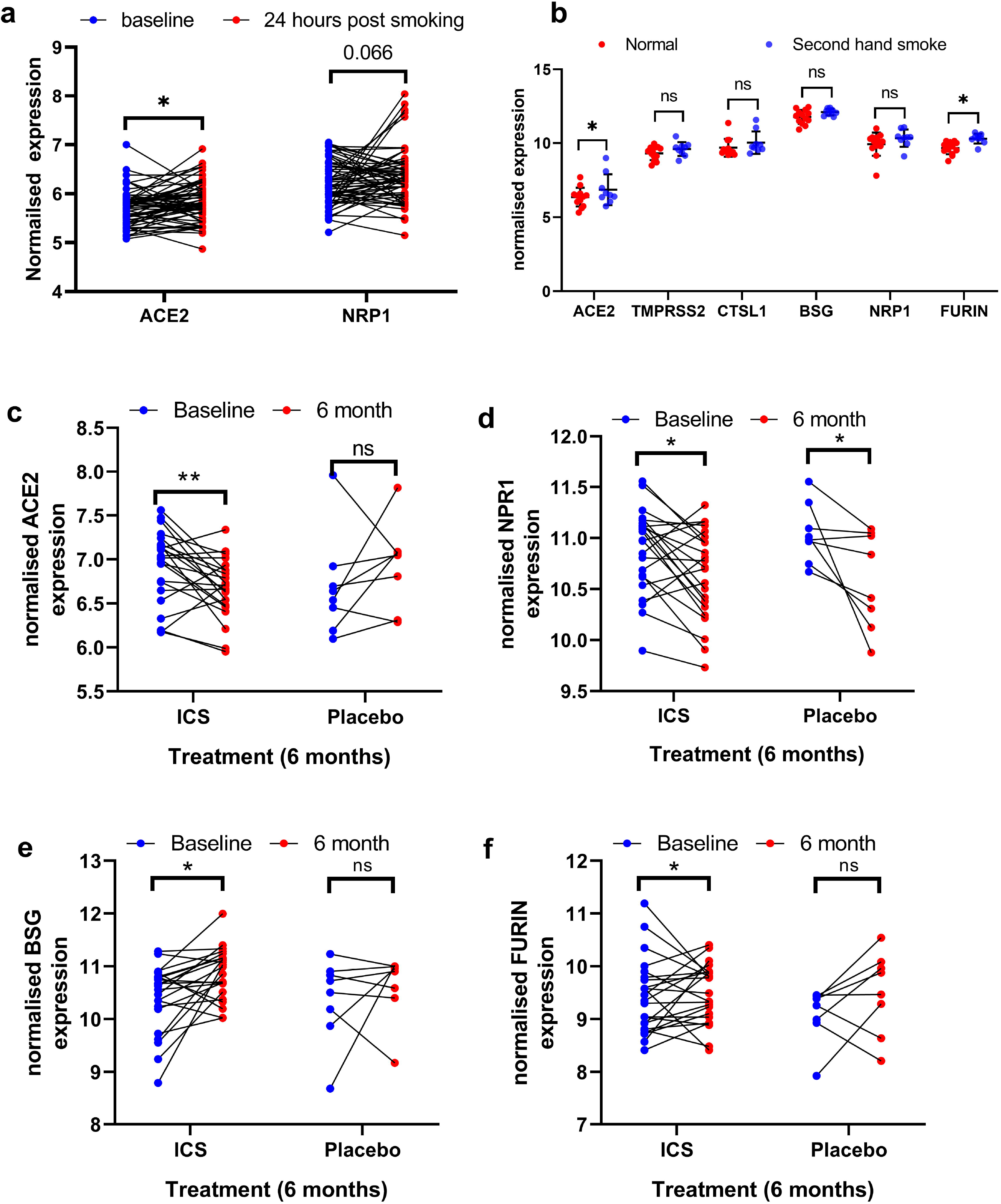
Influence of ICS and acute/ second hand smoke exposure on COVID related genes. The effect of acute smoke exposure on a) ACE2 and NRP1 in bronchial brushings 24 hours after smoking and not smoking 3 cigarettes. b) The influence of second hand smoke in children of COVID related genes in bronchial biopsies. The influence of 6 month ICS and Placebo compared to baseline from bronchial biopsies of COPD patients, c) ACE2, d) NRP1 e) BSG and f) FURIN. *=p<0.05

### Inhaled corticosteroid treatment decreases ACE2 expression in bronchial biopsies

Previous cross-sectional studies have provided suggestive evidence that inhaled corticosteroids (ICS) decrease ACE2 receptor expression in sputum cells in asthmatics^26,27^. To determine whether ICS can influence ACE2 and other SARS-CoV-2 cell entry genes in bronchial biopsies, we investigated the influence of 6- months treatment with ICS with or without added Long-Acting Beta-Agonists (LABA) on longitudinal bronchial biopsy samples in steroid naive COPD patients. We found that ACE2 was significantly decreased by ICS +/- LABA treatment (p=0.009), irrespective of smoking status, while BSG and FURIN were increased (p=0.012 and p=0.046, respectively, Table 7). In addition, NRP1 was also significantly decreased (p=0.009), however, we found a significant effect with the same direction in the placebo group indicating a placebo effect on expression of this gene (Figure 4C-F). Interestingly these changes were not associated with relative cell proportions (data not shown).

**Table 7.**
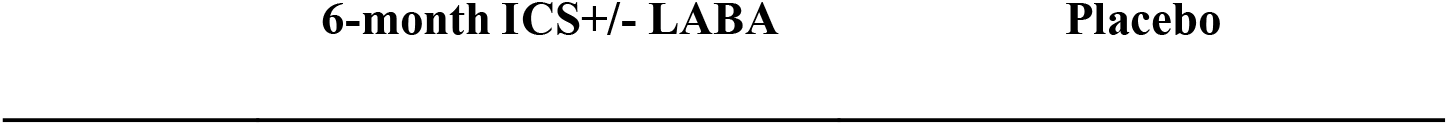
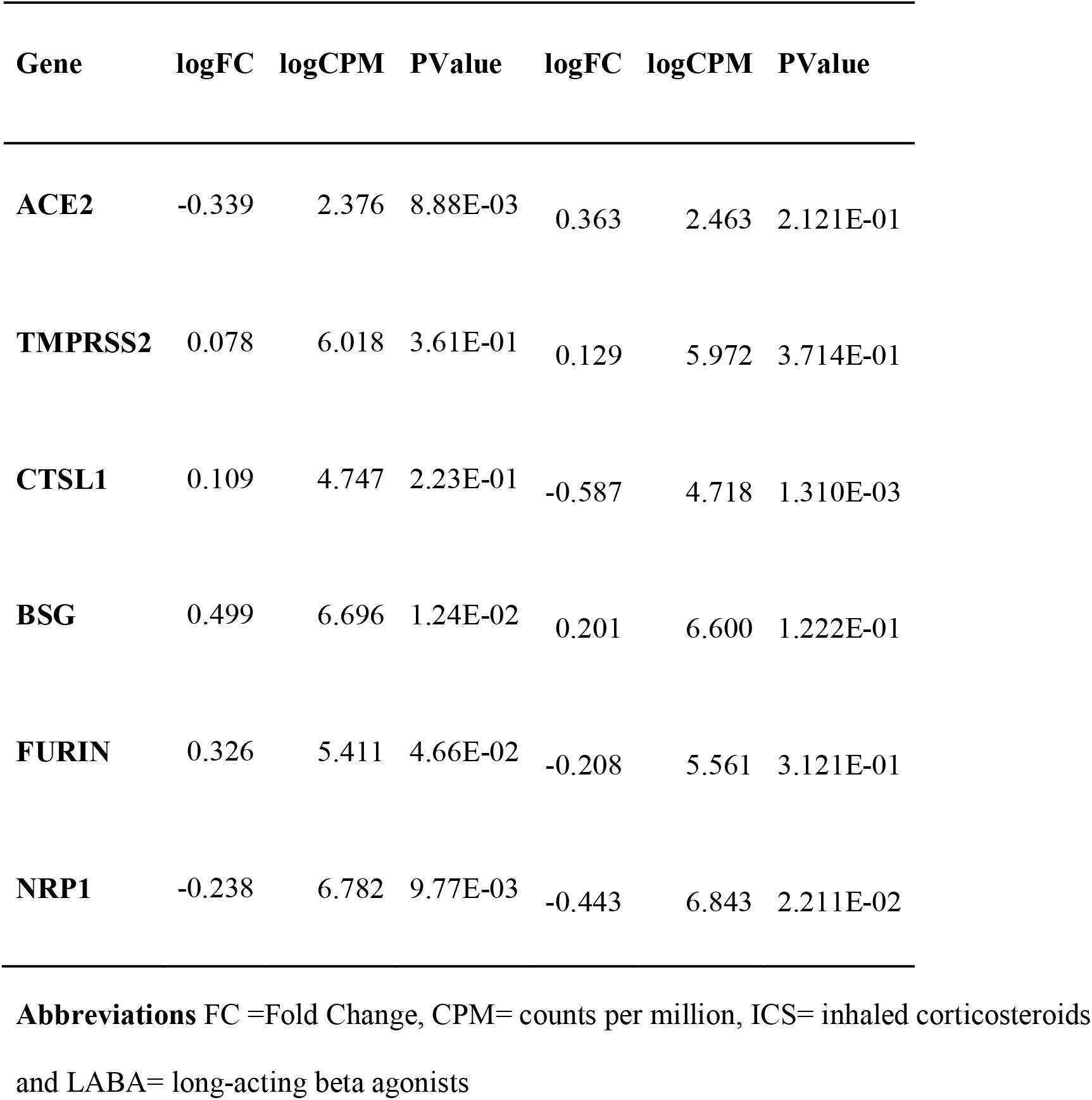
Influence of ICS+/-LABA on expression of COVID related genes in bronchial biopsies.

### The presence of asthma and COPD does not influence the expression of COVID-19 related genes

It has been a concern that the presence of chronic lung disease may increase of the susceptibility for COVID-19 via changes in SARS-CoV-2 cell entry gene expression. Therefore, we tested the influence of disease (COPD or mild Asthma) on gene expression in nasal and bronchial biopsies. The presence of Asthma was investigated in two datasets with nasal tissue (PIAMA and OLIVA/NORM) and one dataset with bronchial tissue (INDURAIN). No effect of asthma was found on the expression levels of SARS-CoV-2 cell entry genes in single-cohort analyses or in meta-analysis Next, we investigated the effect of COPD status (stratified into “none”, “mild”, “moderate” and “severe” categories) on the expression of the 6 COVID related genes both in nasal (n=343) and bronchial (n=184) samples from the CRUCKPAP cohort. We found decreased expression of *TMPRSS2* in nasal samples (p=1.95E-03, logFC=-0.105) and of *CTSL* in bronchial samples (p=8.06 E-04, logFC=-0.307) in relation to the severity of COPD.

### Presence of SNPs do not influence the expression of ACE2 and other COVID related genes in the nose and the bronchus

A number of recent publications report genetic polymorphisms that are associated with SARS-CoV-2 cell entry gene expression levels. We were able to perform eQTL analyses in both bronchial and nasal samples. In nasal RNA-Seq datasets from NORM (n=93), CRUKPAP dataset (n=339) and PIAMA (n=303) we identified an eQTL for *NRP1* (rs10763962, beta=0.378, pvalue=1.03E-06, FDR= 0.002) in only a single cohort (Table S4), while no significant eQTLs were identified for all other COVID related genes in nasal brushing, in contrast to a previous publication ^28^. In bronchial brushings from NORM (n=150) and CRUKPAP (n=215), a single eQTL was identified for *FURIN* (rs401549, beta=0.423, pvalue=2.48E-05, FDR=0.049), only in a single cohort, while no significant eQTLs were identified for all other genes.

### DNA methylation modulates the expression of genes associated with SARS-CoV-2 entry in the upper and lower airways

To investigate the possible effect of DNA methylation on genes associated with SARS-CoV-2 cell entry, an expression Quantitative Trait Methylation (eQTM) analysis was run on nasal and bronchial samples. Nasal eQTM was run on PIAMA samples (n=245), for which CTSL,BSG, NRP1, FURIN and TMPRSS2 expression were associated with CpG methylation levels (Table S5). Bronchial eQTMs were analyzed in INDURAIN (n=169). Here, we identified 143 eQTMs with all the different SARS-CoV-2 related genes hainge one or more significant methylation sites associated with their gene expression levels (Figure 5A-C, Table S5). The nasal eQTMs were influenced by age and sex, while smoking status had no effect (Table S6). The bronchial eQTMs for *TMPRSS2* were found to be associated with smoking status and age (Table S7). Interestingly, three of the five *ACE2* eQTMs were associated with sex, however the most significant *ACE2* eQTM was not associated with any clinical variable. Finally we associated the identified methylation sites with cellular deconvolution profiles to determine if they were cell-type driven. Here we found that the majority of the top eQTMs were strongly negatively associated with ciliated cell proportions (Figure 5D). *ACE2* was found to be associated with 6 CpG sites with 2 sitting in the promoter region of the adjacent gene *TMEM27* in bronchial biopsies (Figure 5E). Direct correlation between *ACE2* and *TMEM27* showed a significant positive regulation (Figure 5F). The same CpG site was found to be an eQTM for *TMEM27* (cg20473453) (Figure 5G), indicating a possible co-regulation of *ACE2* and *TMEM27* genes. This finding identifies a possible new mechanism for the regulation of ACE2.

**Figure 5.**
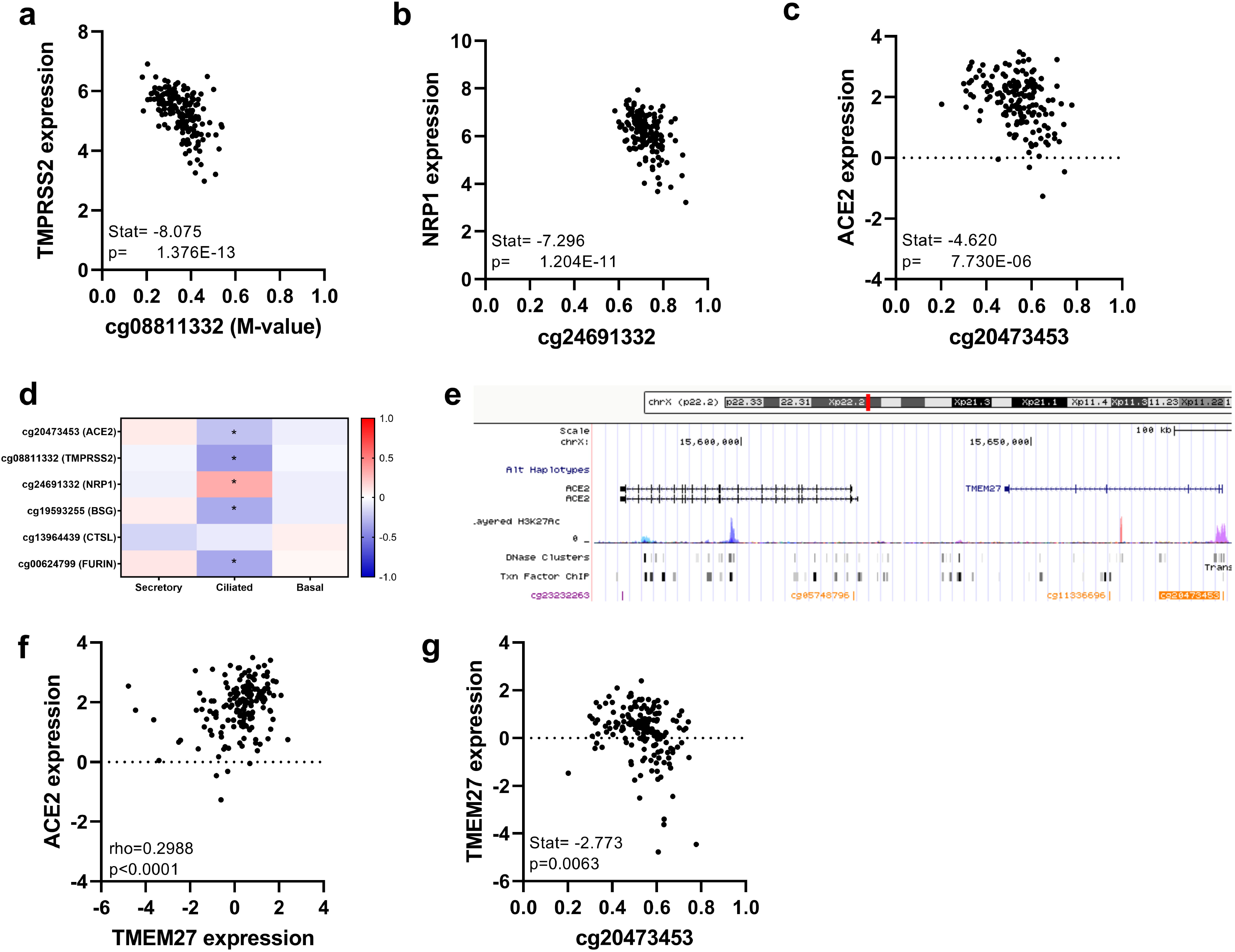
Influence of epigenetics on the expression of COVID related genes. Top eQTM for a) TMPRSS2, b) NRP1 and c) ACE2. d) Correlation matrix of TOP CpG sites and deconvolution of structural cells. e) Diagram of the top CpG site associated with ACE2 expression. f) Correlation of ACE2 and TMEM27. g) EQTM for TMEM27 and the top CpG site associated with ACE2 expression.

## Discussion

The current study investigated the expression of genes required for COVID-19 entry *(ACE2, TMPRSS2, BSG*, *FURIN, NRP1* and *CTSL*) in cells of the upper and lower airways and provided evidence of molecular mechanisms that modify their expression in these compartments. In matched samples from the upper and lower airways, we observed an overall higher expression of these SARS-CoV-2 cell entry genes in the nose. Current smoking was found to be associated with higher gene expression, but only in the lower airways. Both baseline expression in nose and smoke-enhanced expression in the bronchi was found to be associated with goblet cell proportions estimated using cell-type deconvolution. We did not observe an influence of chronic respiratory diseases on COVID related gene expression, but inhaled corticosteroids were found to significantly decrease *ACE2* after 6 months treatment in steroid naive COPD patients. As ICS is a common therapy for asthma and COPD the lack of disease association may be confounded by their current treatment. Finally, we did not observe any significant eQTL effects for the majority of these gene including *ACE2*, in contrast to a recent publication ^28^. However, we do observe that DNA methylation was strongly associated with expression levels of *ACE2, TMPRSS2* and *CTSL*. Furthermore, we find that *ACE2* is co-expressed with an adjacent gene *TMEM27* and is highly associated with a methylation site in the *TMEM27* promoter indicating a possible co-regulation or interaction of the two genes that is independent of smoking status.

The results from our cell-type deconvolution across several independent cohorts reproducibly find that SARS-CoV-2 cell entry genes are most strongly associated with mucus producing cell proportions while no association with ciliated cells, which is in line with single cell RNA-sequencing results reporting expression of these receptors in secretory cells^24,29^. A key observation in this study was the lack of association of smoking with the expression of ACE2 and other COVID related genes in the nose. The explanation for this finding is twofold 1) the selective expression of ACE2 and the other COVID related genes in goblet cells, indicated a possible lack of goblet cell hyperplasia in the nose of current smokers; 2) as the expression of these genes is much higher in nose than in bronchus there is possibly a plateau effect with the level of expression. Our current study highlights the importance of sampling of different tissues. Notwithstanding the relevance of goblet cell metaplasia for induction of ACE2 gene expression by smoke, acute exposure is sufficient to increase ACE2 expression within 24 hours in vivo, indicating that increased expression within the epithelial cells might also contribute.

While gene expression data from our study as well as a number of single-cell and bulk RNA-sequencing analyses indicate expression of SARS-CoV2 entry genes in the secretory cells of the airway epithelium (club and goblet cells), recent data from IHC stainings for ACE2 and analysis of the presence of viral RNA during SARS-CoV-2 infections seem to indicate that ciliated cells, rather than secretory cells express ACE2 protein and are infected by the virus, whereas this has been observed for secretory cells in some studies to a lesser extent^30^. The infection of lower airway epithelial cells as observed by^21^ might be explained by an induction of ACE2 expression in these cells as part of an interferon-induced response (Ziegler et al) after initial upper airway infection. Our data indicate baseline expression levels, and these might rapidly change upon initial infection with SARS-CoV-2 that is thought to occur most likely in the upper, not the lower airways ^20^. Also, such differences in RNA and protein expression levels between cell types might indicate a difference in turnover of the protein, leading to higher RNA expression levels in cells with high protein turnover and less in those with more stable protein. Further studies will need to reveal the relative contribution of secretory and ciliated cells in the upper airways during initial infection.

In the current study, we did not find a relationship between disease status (mild to moderate asthma and COPD) and SARS-CoV-2 cell entry genes. However, a recent study found that ACE2 was increased in severe asthma as compared to healthy, mild and moderate asthma^31^, indicating that disease severity may play an important role in COVID-19 infection, while a recent epidemiological study showed that COPD patients with COVID-19 who were current smokers were at greater risk of severe complications and higher mortality rate, compared to COPD patients who were former and never smokers^32^.

Of clear concern in the current study is the finding that second hand smoke exposure early in life is sufficient to increase the expression of ACE2 and FURIN in the airways of children. This result suggests that children living with smoking parents may be more susceptible to SARS-CoV-2 infection, in addition to the concern of increased exposure due to second hand smoke particles and droplets containing viral loads once their smoking family member becomes infected. It has already been well established that children exposed to secondhand smoke are more susceptible to common respiratory infections and are predisposed to developing chronic inflammatory airways diseases ^33,34^ Although COVID-19 symptoms are usually mild in children, our findings underline the importance of smoking cessation in caregivers of young children.

Corticosteroids play an important role in the suppression of inflammation in chronic inflammatory diseases such as asthma and COPD. Previous cross-sectional studies have provided evidence that inhaled corticosteroids are associated with lower ACE2 receptor expression in sputum samples derived from asthmatic patients^26,27^. We now extend these findings in a longitudinal study by showing that inhaled corticosteroids also decrease ACE2 receptor mRNA expression in bronchial biopsies of patients with COPD. Our findings support the notion that inhaled corticosteroids are unlikely to increase the risk for more severe COVID-19 disease and should be continued in patients with obstructive pulmonary diseases. However we did find the expression level of FURIN and BSG (SARS-CoV-2 alternative receptor) to be increased with ICS treatment.

The methylation results in the current manuscript provide a novel mechanism for the regulation of SARS-CoV-2 cell entry gene expression. Most eQTMs were negatively correlated with ciliated cell proportions, indicating a reduced methylation levels of the corresponding CpG sites, which would possibly lead to an upregulation of the associated genes. This would match protein data, which shows high expression of ACE2 in ciliated cells as discussed above. Furthermore, we found a possible co-regulation or interaction of ACE2 with the promoter for TMEM27 (an adjacent gene).

There were a number of limitations associated with the current study. In the current study we found a number of associations of COVID related genes with epidemiological factors and current treatment at a gene expression level. However, RNA does not always equal protein levels and extensive immunohistochemical stainings for these genes needs to be done to confirm a number of these findings. Furthermore, the current data provides only a snapshot pre-infection, and many of these genes will change rapidly following a COVID-19 infection. The current study provides the first longitudinal evaluation of ACE2 expression in bronchial biopsies on a limited number of samples which deserves expansion in subsequent studies.

In conclusion we find that genes required for COVID-19 entry into cells are highly expressed in the upper airways compared to the lower airways by analysis of matched samples. Both first and second hand smoke exposure were found to be associated with higher expression of genes required for SARS-CoV-2 entry into cells. Methylation was found to influence the expression of a number of these genes in the bronchus and nose, which appeared to be regulated by age, gender and smoking status. Finally ICS were found to decrease the expression of ACE2 expression in bronchial biopsies.

## Data Availability

The curent manuscript involves a number of studies which have been uploaded to GEO and other data sharing websites

## References

1 Chen N, Zhou M, Dong X, et al. Epidemiological and clinical characteristics of 99 cases of 2019 novel coronavirus pneumonia in Wuhan, China: a descriptive study. Lancet 2020; published online Jan 30. DOI:10.1016/S0140-6736(20)30211-7.

2 Zhu N, Zhang D, Wang W, et al. A Novel Coronavirus from Patients with Pneumonia in China, 2019. N Engl J Med 2020; published online Jan 24. DOI:10.1056/NEJMoa2001017.

3 Zhou F, Yu T, Du R, et al. Clinical course and risk factors for mortality of adult inpatients with COVID-19 in Wuhan, China: a retrospective cohort study. Lancet 2020; 395: 1054-62.

4 Wang D, Hu B, Hu C, et al. Clinical Characteristics of 138 Hospitalized Patients With 2019 Novel Coronavirus-Infected Pneumonia in Wuhan, China. JAMA 2020; 323: 1061-9.

5 Berlin DA, Gulick RM, Martinez FJ. Severe Covid-19. N Engl J Med 2020; published online May 15. DOI:10.1056/NEJMcp2009575.

6 Richardson S, Hirsch JS, Narasimhan M, et al. Presenting Characteristics, Comorbidities, and Outcomes Among 5700 Patients Hospitalized With COVID-19 in the New York City Area. JAMA 2020; published online April 22. DOI:10.1001/jama.2020.6775.

7 Mao L, Jin H, Wang M, et al. Neurologic manifestations of hospitalized patients with coronavirus disease 2019 in Wuhan, China [published online April 10, 2020]. JAMA Neurol.

8 Huang C, Wang Y, Li X, et al. Clinical features of patients infected with 2019 novel coronavirus in Wuhan, China. Lancet 2020; published online Jan 24. DOI:10.1016/S0140-6736(20)30183-5.

9 Wu C, Chen X, Cai Y, et al. Risk Factors Associated With Acute Respiratory Distress Syndrome and Death in Patients With Coronavirus Disease 2019 Pneumonia in Wuhan, China. JAMA Intern Med 2020; published online March 13. DOI:10.1001/jamainternmed.2020.0994.

10 Vardavas CI, Nikitara K. COVID-19 and smoking: A systematic review of the evidence. Tob Induc Dis 2020; 18: 20.

11 Hoffmann M, Kleine-Weber H, Krüger N, Müller M, Drosten C, Pöhlmann S. The novel coronavirus 2019 (2019-nCoV) uses the SARS-coronavirus receptor ACE2 and the cellular protease TMPRSS2 for entry into target cells. Molecular Biology. 2020;: 1136.

12 Wang Q, Zhang Y, Wu L, et al. Structural and Functional Basis of SARS-CoV-2 Entry by Using Human ACE2. Cell 2020; 181: 894-904.e9.

13 Wang K, Chen W, Zhou Y-S, et al. SARS-CoV-2 invades host cells via a novel route: CD147-spike protein. bioRxiv. 2020;: 2020.03.14.988345.

14 Daly JL, Simonetti B, Plagaro CA, Williamson MK. Neuropilin-1 is a host factor for SARS-CoV-2 infection. *bioRxiv* 2020. https://www.biorxiv.org/content/10.1101/2020.06.05.134114v1.abstract.

15 Cantuti-Castelvetri L, Ojha R, Pedro LD, Djannatian M. Neuropilin-1 facilitates SARS-CoV-2 cell entry and provides a possible pathway into the central nervous system. *bioRxiv* 2020. https://www.biorxiv.org/content/10.1101/2020.06.07.137802v1.abstract.

16 Millet JK, Whittaker GR. Host cell proteases: Critical determinants of coronavirus tropism and pathogenesis. Virus Res 2015; 202: 120-34.

17 Ellinghaus D, Degenhardt F, Bujanda L, et al. Genomewide Association Study of Severe Covid-19 with Respiratory Failure. N Engl J Med 2020; published online June 17. DOI:10.1056/NEJMoa2020283.

18 Regev A, Teichmann SA, Lander ES, et al. The Human Cell Atlas. Elife 2017; 6. DOI:10.7554/eLife.27041.

19 Lukassen S, Chua RL, Trefzer T, et al. SARS-CoV-2 receptor ACE2 and TMPRSS2 are primarily expressed in bronchial transient secretory cells. EMBO J 2020; 39: e105114.

20 Sungnak W, Huang N, Bécavin C, et al. SARS-CoV-2 entry factors are highly expressed in nasal epithelial cells together with innate immune genes. Nat Med 2020; 26: 681-7.

21 Ziegler CGK, Allon SJ, Nyquist SK, et al. SARS-CoV-2 Receptor ACE2 Is an Interferon-Stimulated Gene in Human Airway Epithelial Cells and Is Detected in Specific Cell Subsets across Tissues. Cell 2020; 181: 1016-35.e19.

22 Hikmet F, Méar L, Edvinsson Å, Micke P, Uhlén M, Lindskog C. The protein expression profile of ACE2 in human tissues. Mol Syst Biol 2020; 16: e9610.

23 Vieira Braga FA, Kar G, Berg M, et al. A cellular census of human lungs identifies novel cell states in health and in asthma. Nat Med 2019; 25: 1153-63.

24 Muus C, Luecken MD, Eraslan G, Waghray A. Integrated analyses of single-cell atlases reveal age, gender, and smoking status associations with cell type-specific expression of mediators of SARS-CoV-2 …. BioRxiv 2020. https://www.biorxiv.org/content/10.1101/2020.04.19.049254v1.full-text.

25 Imkamp K, Bernal V, Grzegorzcyk M, et al. Gene network approach reveals co-expression patterns in nasal and bronchial epithelium. Sci Rep 2019; 9: 15835.

26 Finney LJ, Glanville N, Farne H, Aniscenko J. Inhaled corticosteroids downregulate the SARS-CoV-2 receptor ACE2 in COPD through suppression of type I interferon. bioRxiv 2020. https://www.biorxiv.org/content/10.1101/2020.06.13.149039v1.abstract.

27 Peters MC, Sajuthi S, Deford P, et al. COVID-19-related Genes in Sputum Cells in Asthma. Relationship to Demographic Features and Corticosteroids. Am J Respir Crit Care Med 2020; 202: 83-90.

28 Cao Y, Li L, Feng Z, et al. Comparative genetic analysis of the novel coronavirus (2019-nCoV/SARS-CoV-2) receptor ACE2 in different populations. Cell Discov 2020; 6: 11.

29 Smith JC, Sheltzer JM. Cigarette smoke triggers the expansion of a subpopulation of respiratory epithelial cells that express the SARS-CoV-2 receptor ACE2. bioRxiv. 2020. preprint) [CrossRef][Google Scholar] 2020.

30 Mulay A, Konda B, Garcia G, et al. SARS-CoV-2 infection of primary human lung epithelium for COVID-19 modeling and drug discovery. *bioRxiv* 2020; published online June 29. DOI:10.1101/2020.06.29.174623.

31 Kermani N, Song W-J, Lunt A, et al. Airway expression of SARS-CoV-2 receptor, ACE2, and proteases, TMPRSS2 and furin, in severe asthma. medRxiv 2020. https://www.medrxiv.org/content/10.1101/2020.06.29.20142091v1.abstract.

32 Alqahtani JS, Oyelade T, Aldhahir AM, et al. Prevalence, Severity and Mortality associated with COPD and Smoking in patients with COVID-19: A Rapid Systematic Review and Meta-Analysis. PLoS One 2020; 15: e0233147.

33 Lewis SA, Antoniak M, Venn AJ, et al. Secondhand Smoke, Dietary Fruit Intake, Road Traffic Exposures, and the Prevalence of Asthma: A Cross-Sectional Study in Young Children. American Journal of Epidemiology. 2005; 161: 406-11.

34 Kott KS, Salt BH, McDonald RJ, Jhawar S, Bric JM, Joad JP. Effect of secondhand cigarette smoke, RSV bronchiolitis and parental asthma on urinary cysteinyl LTE4. Pediatr Pulmonol 2008; 43: 760-6.

